# Deep AI model for autism detection using naturalistic behavioral videos

**DOI:** 10.1101/2025.10.01.25336912

**Authors:** Guikun Chen, Wei Zhou, Lingli Zhang, Yiting Ji, Qianlong Zhang, Tai Ren, Hangyu Tan, Jingyu Chen, Keyi Liu, Xiaoguang Song, Sikai Huang, Lingqi Gu, Jiayang Liu, Hui Wang, Guanghui Sui, Yujia Wang, Xiaolei Han, Wenguan Wang, Fei Li

**Author notes:** These authors contributed equally to this work (Guikun Chen, Wei Zhou, Lingli Zhang, Yiting Ji).

## Abstract

Profound heterogeneity in Autism Spectrum Disorder (ASD) complicates diagnosis and the development of effective treatments. In healthcare systems with limited specialist resources, the need for rapid and accessible screening tools is particularly urgent. In the present study, we developed an objective, scalable pipeline that pairs a simple two-minute video recording of a child’s naturalistic behavior with a deep AI model for autism detection, representing the first such rapid, scalable framework. By analyzing a rich spectrum of children’s responses to social stimuli, such as subtle behavioral patterns, gaze dynamics, facial morphology, and dynamic facial complexity, the deep AI model provides powerful support for clinical workflows, demonstrating high accuracy in identifying ASD risk across diverse internal and external test cohorts, irrespective of sex, age, or cognitive function. Furthermore, a series of comprehensive analyses confirmed the model’s clinical relevance and revealed its capacity to objectively stratify ASD heterogeneity into neurobiologically distinct subtypes. This work establishes a highly efficient and objective framework for large-scale screening, providing a data-driven foundation to stratify heterogeneity and paving the way for the future development of targeted interventions.

## 1 Introduction

Autism Spectrum Disorder (ASD) is a neurodevelopmental condition characterized by a wide range of social communication abilities, patterns of restricted and repetitive behaviors, and diverse cognitive profiles. This heterogeneity makes autism a significant challenge for clinical practice and research. An accurate and timely diagnosis is critical for individuals to access appropriate support and for families to understand their child’s needs [1, 2]. However, the profound heterogeneity complicates the diagnostic process, making it difficult to apply a uniform standard across all individuals. Furthermore, this same variability presents a major obstacle to developing broadly effective treatments and identifying distinct biological subgroups [3–5]. A central goal in autism research is therefore to develop objective methods that can systematically parse this heterogeneity, which is essential for improving diagnosis and paving the way for more personalized and effective interventions [3].

The importance of early and accurate diagnosis is undisputed, as interventions initiated within critical neurodevelopmental windows, such as the first 36 months of life, can significantly improve long-term outcomes [6]. Current diagnostic practices rely on expert observation of a child’s behavior and developmental history. Standardized tools such as the Autism Diagnostic Observation Schedule (ADOS) and the Autism Diagnostic Interview-Revised (ADI-R) are commonly used to aid in this process. Recently, the field has advanced towards objective behavioral markers using technologies like computer vision analysis [7, 8]. Concretely, mainstream assessment tools [7], which demonstrated high accuracy in research settings, rely on elaborate data collection protocols. These often involve a sequence of structured tasks, such as requiring the child to watch specific videos, interact with touch-screen applications, or respond to directed social prompts, collectively demanding approximately 10 minutes or more of sustained and varied engagement.

The complexity of these multi-step protocols creates a significant practical barrier to their widespread use. Such lengthy and varied tasks are often difficult for young children to complete, particularly those with emerging traits of autism who may struggle with sustained attention, complex instructions, or cooperative participation. This makes these protocols largely infeasible for scalable implementation in busy, real-world environments like primary care or community centers. This challenge is magnified by the limited availability of specialist clinicians, which already creates diagnostic delays [9–12]. The reliance on complex procedures and specialist resources limits the utility of current methods for widespread screening and hinders progress toward equitable and accessible diagnostic support.

To address the data acquisition challenges, we developed a new pipeline that pairs a simple, two-minute video recording of a child’s behavior with a deep AI model for autism detection. In our approach, the child only needs to passively watch a short video, removing the need for complex instructions or multiple tasks. The simplicity of the data acquisition process makes it practical for integration into routine pediatric visits, creating more opportunities for early identification and access to timely support. Our model achieves high accuracy in identifying ASD risk across diverse internal and external test cohorts, irrespective of sex, age, or cognitive function. Together, this work represents a rapid, objective, and scalable screening tool that can be used to enhance clinical workflows in diverse healthcare settings.

Beyond practical barriers in data acquisition, current computational methods for autism detection face methodological limitations. Current AI-driven methods often rely on traditional machine learning models (e.g., XGBoost [7, 13]) that operate on hand-crafted features, where researchers pre-select specific behaviors for analysis. This process can be labor-intensive and may introduce biases based on existing assumptions [14, 15], potentially overlooking the full spectrum of relevant behavioral patterns. Furthermore, these models are typically designed only to detect behavioral differences and do not classify individuals into meaningful subgroups. This focus on detection alone creates a disconnect between observable behavioral phenotypes and the underlying brain characteristics, which hinders mechanistic understanding and the development of targeted interventions.

Our work makes use of an end-to-end deep learning model [16, 17] to overcome these limitations. The model learns relevant features directly from raw video data, minimizing reliance on pre-specified assumptions and enabling the discovery of novel behavioral patterns. Crucially, when applied to data modalities like video analysis of gaze and social engagement [18, 19], these techniques leverage an established theoretical understanding linking atypical visual attention patterns directly to core ASD characteristics [20]. This direct mechanistic link offers a significant advantage in explainability over other AI applications in ASD research, such as those attempting to derive diagnostic signals from data like retinal photographs [21], where the connection to core neurodevelopmental processes is currently less evident. Furthermore, we conducted a series of comprehensive analyses to confirm the model’s clinical relevance. Experiments demonstrate that the model can stratify the heterogeneity of ASD into distinct subtypes [22, 23] based on the behavioral patterns it learns. These data-driven subgroups were subsequently validated against clinical assessments and neuroimaging data, revealing that they correspond to neurobiologically distinct profiles. This work establishes a framework that not only improves the efficiency of detection but also connects objective behavioral measures to underlying biology, paving the way for deeper understanding of autism and the development of stratified interventions.

## 2 Results

### Diagnostic accuracy of foundational AI models for autism detection

To assess the capability of foundational AI models in detecting ASD from behavioral video data (Fig. 1), we employed an end-to-end deep learning approach centered on VideoMAE [16], a foundational video AI model. VideoMAE, which is pre-trained on the large-scale Kinetics-400 dataset and subsequently fine-tuned on our cohort’s naturalistic video recordings, was evaluated on an independent internal test (discovery) set (*n* = 126, Tables 1 and 2) to provide an unbiased assessment of its performance, with a validation set used for model selection. On this internal test set, the model demonstrated robust diagnostic capability (Table 3), achieving a primary endpoint Receiver Operating Characteristic Area Under the Curve (ROC AUC) of 0.937 (confidence interval (CI) (0.89-0.97)). This high AUC indicates the model’s strong ability to discriminate between children with ASD and those without across various decision thresholds. At the optimal operating point determined by maximizing the Youden index, the model yielded an accuracy of 88.2% (s.d. = 2.7%), a sensitivity of 86.8% (s.d. = 7.5%), a specificity of 89.7% (s.d. = 6.9%), a positive predictive value (PPV) of 90.0% (s.d. = 6.5%), and a negative predictive value (NPV) of 87.6% (s.d. = 6.0%). To rigorously test the model’s real-world generalizability, we performed external validation on an independent cohort from a different clinical network, thereby introducing heterogeneity in patient demographics, clinical presentation, and recording environments. As shown in the bottom row of Table 3, the model achieved a ROC AUC of 0.858 on this challenging external dataset. While this, as anticipated, represents a moderation in performance compared to the validation and discovery set, it demonstrates substantial diagnostic utility without any site-specific fine-tuning.

**Fig. 1:**
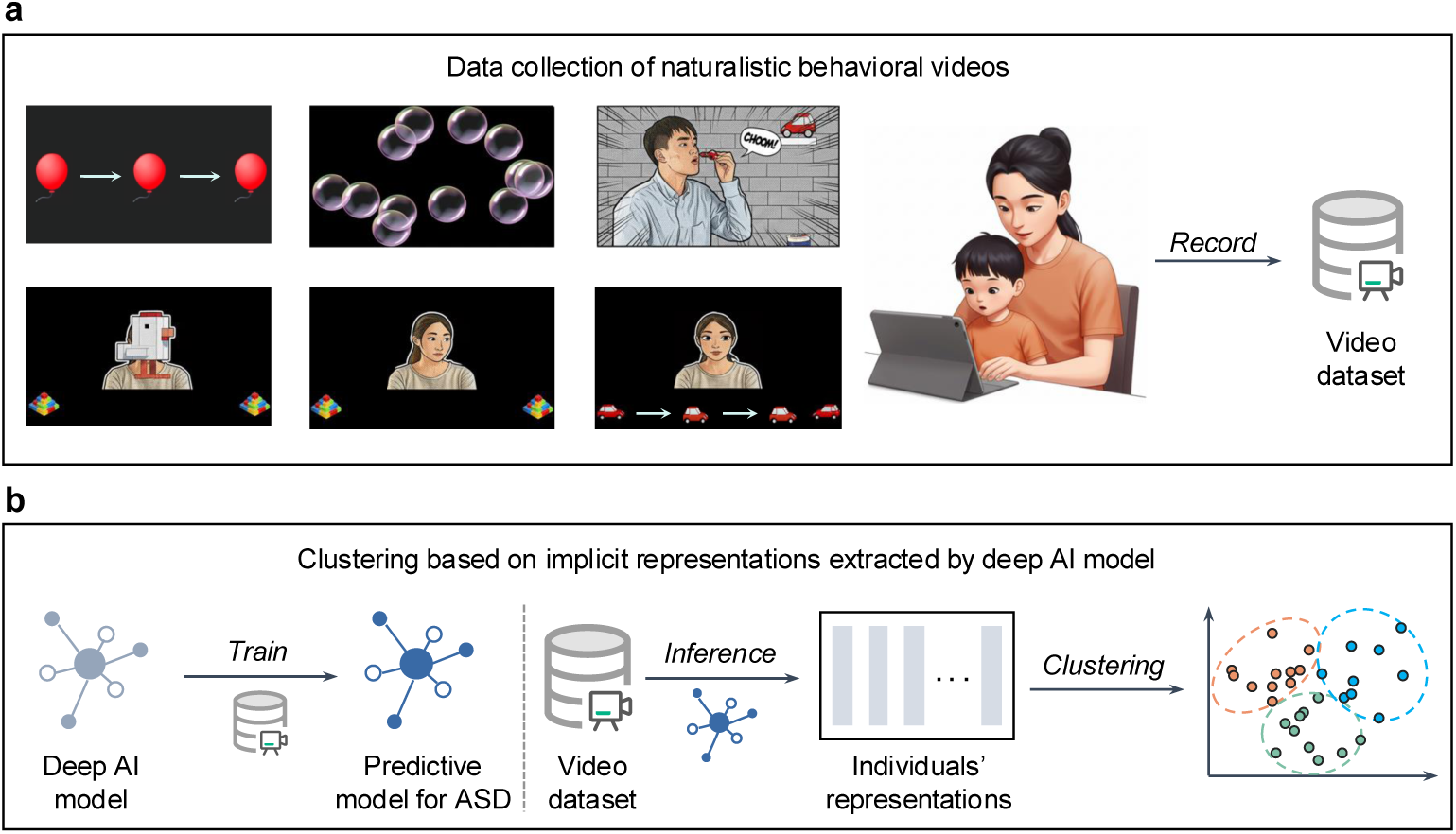
Overview of this study from data collection to automatic clinically actionable subtype identification. **a,** Probing social-communicative behavior with video stimuli. Children were recorded viewing two brief videos designed to elicit key ASD-related behaviors. Stimuli assessed social preference (social vs. non-social targets) and joint attention (both responding joint attention and initiating joint attention). The entire behavioral response was captured in a 2-minute video recording using a standard mobile device. **b**, Clustering learned representations to reveal ASD subtypes. A foundational AI model, fine-tuned for ASD classification, was used to transform each child’s 2-minute behavioral video into a learned feature representation. Unsupervised clustering of these representations stratified the ASD cohort into distinct subtypes, which were subsequently validated against clinical and neurobiological data.

**Table 1:**
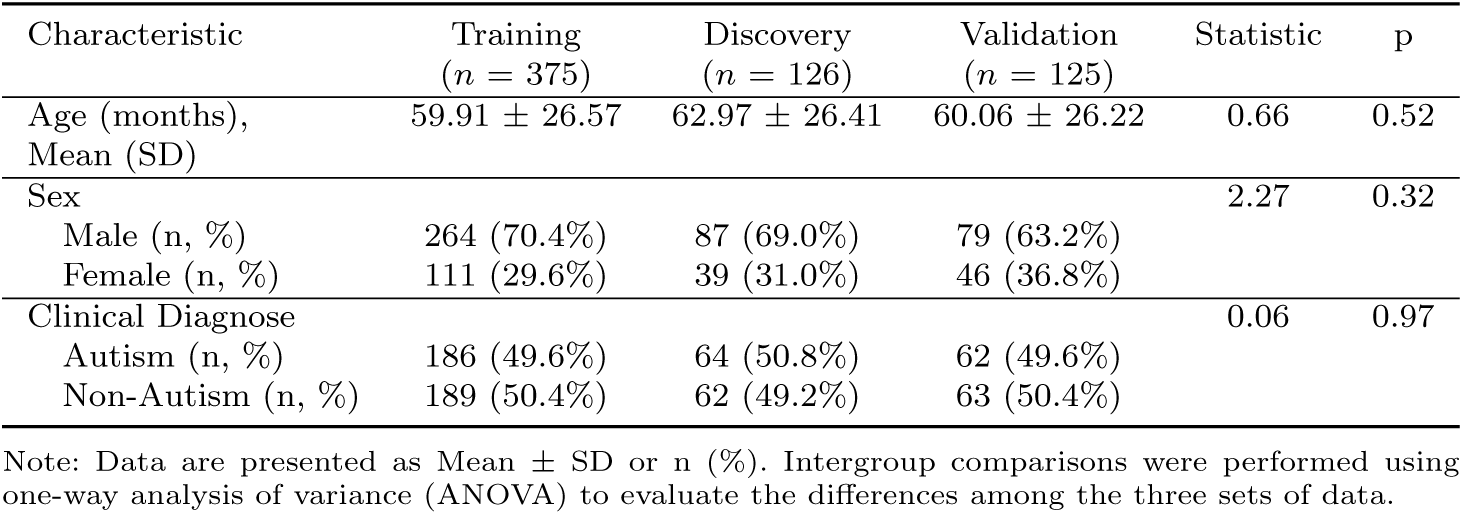
Demographic and clinical features of the training, discovery (test), and validation sets.

**Table 2:** Clinical characteristic of training, discovery (test), and validation sets for autism.

|  | Training<br>(*n* = 186) | Discovery<br>(*n* = 64) | Validation<br>(*n* = 62) | Statistic | p |
| --- | --- | --- | --- | --- | --- |
| CARS total score | 36.28 ± 3.77<br>(*n* = 179) | 36.54 ± 3.99<br>(*n* = 60) | 35.93 ± 3.54<br>(*n* = 61) | 0.40 | 0.67 |
| SRS total score | 84.22 ± 25.45<br>(*n* = 161) | 90.49 ± 25.50<br>(*n* = 53) | 80.58 ± 27.26<br>(*n* = 52) | 2.02 | 0.13 |
| ADOS total score | 16.31 ± 3.73<br>(*n* = 182) | 16.66 ± 3.83<br>(*n* = 62) | 16.31 ± 3.83<br>(*n* = 61) | 0.21 | 0.81 |
| Comorbidity |  |  |  | 0.48 | 0.79 |
| With DD/ID | 118 (63.4%) | 41 (64.1%) | 43 (69.4%) |  |  |
| Without DD/ID | 55 (29.6%) | 19 (29.7%) | 16 (25.8%) |  |  |

Note: Data are presented as Mean ± SD or n (%). Intergroup comparisons were performed using one-way analysis of variance (ANOVA) to evaluate the differences among the three sets of data. The childhood autism rating scale (CARS) and the autism diagnostic observation schedule (ADOS) are standardized, clinician-administered instruments for assessing autism spectrum disorder. Complementing these tools, the social responsiveness scale (SRS) quantifies the full spectrum of social-communication impairments, providing a dimensional measure of symptom severity. DD/ID (developmental delay/intellectual disability) was defined by clinical assessment. Patients were diagnosed with DD/ID if their developmental quotient (DQ) was < 75 on the Gesell Development Scales or their intelligence quotient (IQ) was < 70 on the WISC-R or WPPSI (Wechsler Intelligence Scale for children).

**Table 3:**
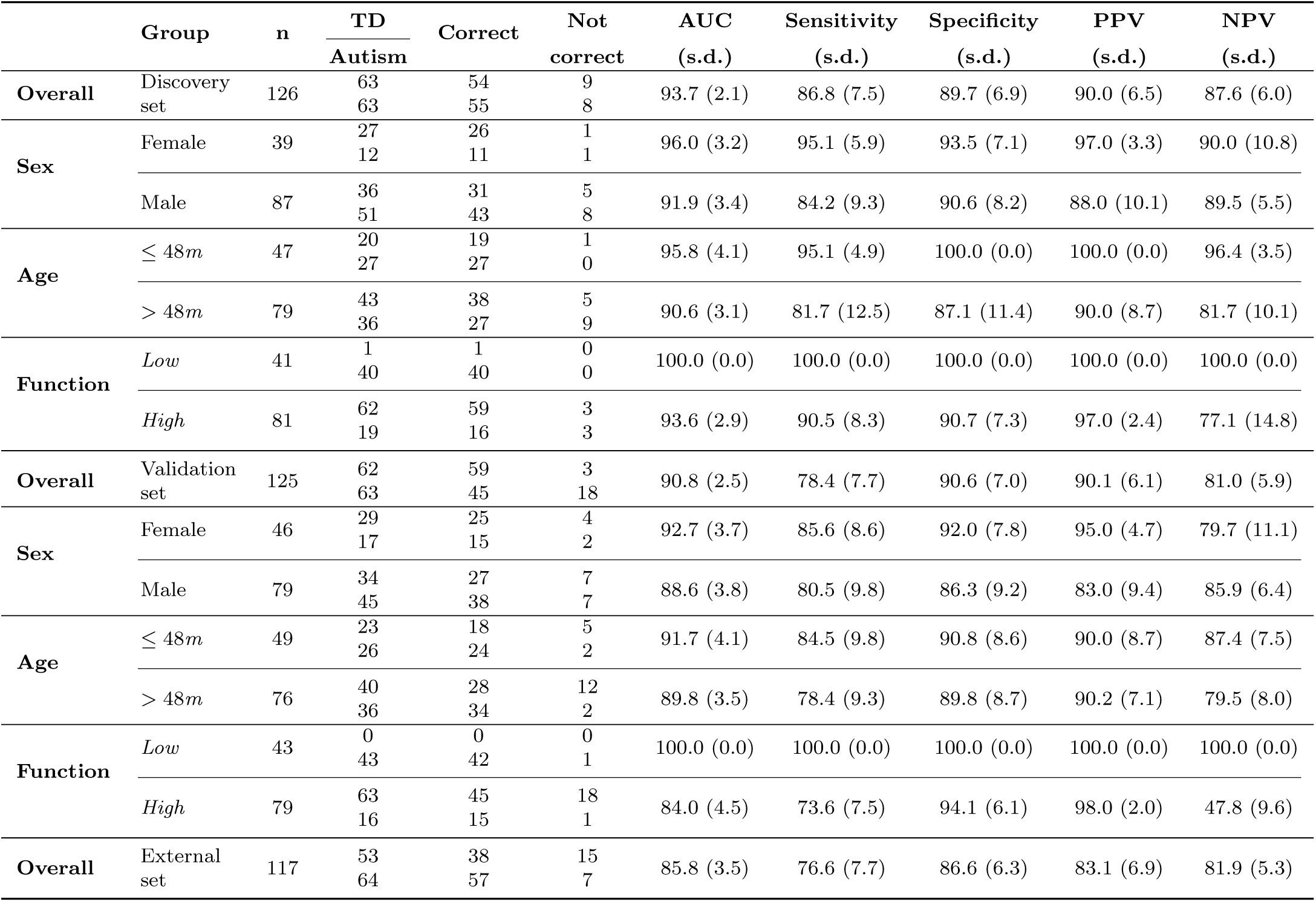
Participant characteristics and model performance metrics (95% CI) in the discovery, validation, and external sets. Standard deviation is shown in parentheses. Here, PPV, NPV, and TD represent positive predictive value, negative predictive value, and typical developing children, respectively.

This successful generalization highlights a fundamental advantage of our end-to-end approach. Unlike models reliant on handcrafted features, which are brittle to variations in how pre-defined behaviors manifest across different settings, our foundational model learns relevant spatiotemporal patterns directly from raw video data. Its ability to maintain high performance when confronted with this real-world variability is a critical finding, suggesting it overcomes a key hurdle for clinical translation. These results therefore underscore the promise of foundational AI to deliver scalable and objective diagnostic aids that are robust enough for deployment across diverse healthcare settings.

### Diagnostic accuracy for subgroups

A critical aspect of any diagnostic tool is its consistent performance across diverse populations. We therefore evaluated the model’s accuracy within key demographic and clinical subgroups using both the discovery and validation sets. The results are summarized in Table 3.

Across both discovery and validation cohorts, the model maintained robust performance across sexes. In the discovery set, the model achieved an AUC of 0.960 (s.d. = 3.2) for females (*n* = 39) and 0.919 (s.d. = 3.4) for males (*n* = 87). Correspondingly, in the validation set, AUCs were 0.927 (s.d. = 3.7) for females (*n* = 46) and 0.886 (s.d. = 3.8) for males (*n* = 79). While performance metrics (e.g., sensitivity, specificity) were numerically slightly higher in females across both sets, the overlapping standard deviations suggest no statistically significant difference in diagnostic accuracy between sexes.

Given the importance of early detection, we examined performance across age groups (*≤*48 months vs. *>*48 months). The model consistently demonstrated strong diagnostic accuracy for both younger and older children. In the discovery set, children aged *≤*48 months (*n* = 47) showed an AUC of 0.958 (s.d. = 4.1). For children older than 48 months (*n* = 79), the AUC remained high at 0.906 (s.d. = 3.1). Similar trends were observed in the validation set, with AUCs of 0.917 (s.d. = 4.1) for the younger cohort (*n* = 49) and 0.898 (s.d. = 3.5) for the older cohort (*n* = 76). These findings underscore the model’s utility across a wide age range relevant for autism diagnosis. We also assessed diagnostic accuracy across different cognitive levels (low-functioning vs. high-functioning). For the low-functioning group (discovery *n* = 41, validation *n* = 43), the model showed high or even perfect performance. This is largely attributable to the small sample sizes and the absence of typical developing children in this subgroup, resulting in a dataset primarily composed of individuals with autism. For high-functioning participants (discovery *n* = 81, validation *n* = 79), the model maintained strong performance, with AUCs of 0.936 (s.d. = 2.9) in the discovery set and 0.840 (s.d. = 4.5) in the validation set. These results demonstrate the model’s ability to detect autism across the spectrum of cognitive levels, including more subtle presentations.

Overall, our foundational AI model consistently exhibited robust diagnostic performance across diverse demographic (sex, age) and clinical (cognitive function) subgroups in both discovery and validation datasets. These results suggest the model’s broad applicability and stability across heterogeneous patient populations.

### Correlation between model-derived probability and clinical measures

Beyond binary classification, we assessed the relationships between model-derived probability and a range of clinical assessments within the ASD cohort. We employed both linear regression and partial correlation analyses to ensure the robustness of these associations. Our analysis revealed that the model-derived probability served as a strong and statistically significant predictor for multiple core clinical measures of autism. The linear regression results (Fig. 2a) demonstrate that the model-derived probability can account for a substantial portion of the variance in clinical assessments. The strongest association was observed with the CARS Social Impairment subscale, where the model-derived probability explained 26.3% of the variance in clinical scores (adjusted R^2^=0.263, standardized *β*=0.21, p*<*0.001). This was closely followed by the CARS Total score (adjusted R^2^=0.236, standardized *β*=0.20, p*<*0.001) and the ADOS Total score (adjusted R^2^=0.22, p*<*0.001). Significant predictive relationships were also found for other key domains, e.g., ADOS Social, ADOS Communication, and various SRS subscales (Fig. 2a).

**Fig. 2:**
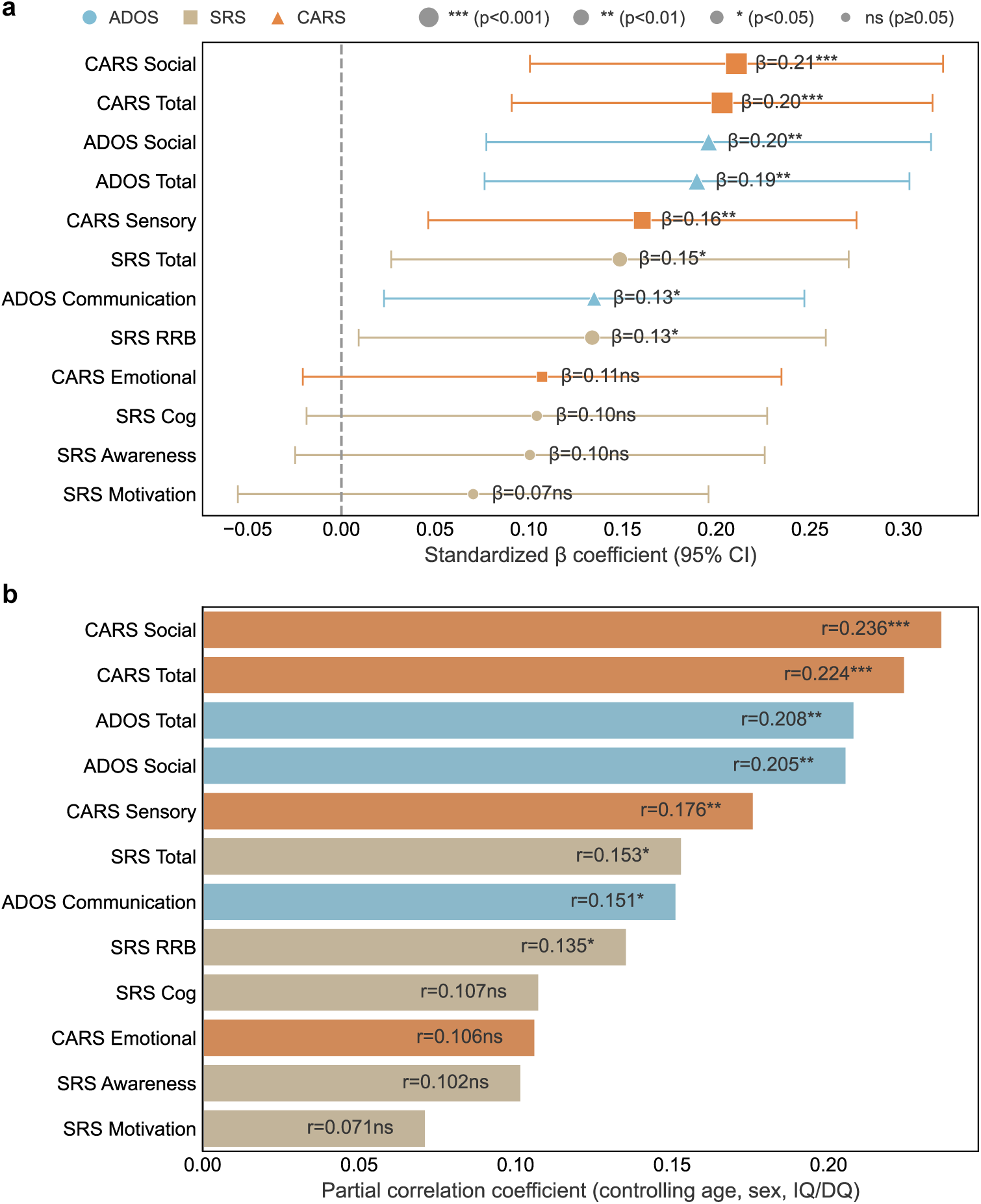
Model-derived probability correlates with established clinical assessments of autism. The figure demonstrates the clinical relevance of the model-derived probability by examining its relationship with established clinical measures in a cohort of individuals with autism spectrum disorder (ASD, *n* = 243). **a**, Forest plot of standardized beta (*β*) coefficients from linear regression models predicting Childhood Autism Rating Scale (CARS), Autism Diagnostic Observation Schedule (ADOS), and Social Responsiveness Scale (SRS) scores using the model-derived probability as the predictor. Error bars represent 95% confidence intervals (CI). **b**, Bar plot showing partial correlation coefficients between the model-derived probability and each clinical measure, adjusted for participant age, sex, and developmental/intelligence quotient (IQ/DQ). This analysis confirms that the associations are independent of key potential confounders. Source data are provided as a Source Data file.

To further strengthen these findings, we conducted a partial correlation analysis to control for potential confounding variables, including participant age, sex, and developmental/intelligence quotient (IQ/DQ). Even after accounting for these factors, the model-derived probability maintained a significant correlation with clinical measures (Fig. 2b). The association with CARS Social Impairment remained the most robust (partial r=0.236, p*<*0.001), followed by CARS Total (partial r=0.224, p*<*0.001) and ADOS Total (partial r=0.208, p*<*0.001).

### Quantification and comparison of core digital behavioral phenotypes

To understand the specific behavioral and facial morphological features that drive the model’s diagnostic classification, we extracted and quantified a comprehensive set of digital phenotypes from the video data and compared them between the ASD and TD groups. These discriminating features spanned three key domains: facial expression dynamics, social attention patterns, and facial morphology. The statistical significance of these group differences is visualized in a lollipop plot (Fig. 3), where each point represents a distinct phenotype. Our analysis revealed numerous phenotypes that significantly differentiated the ASD and TD groups after a stringent False Discovery Rate (FDR) correction for multiple comparisons (adjusted p *<* 0.05).

**Fig. 3:**
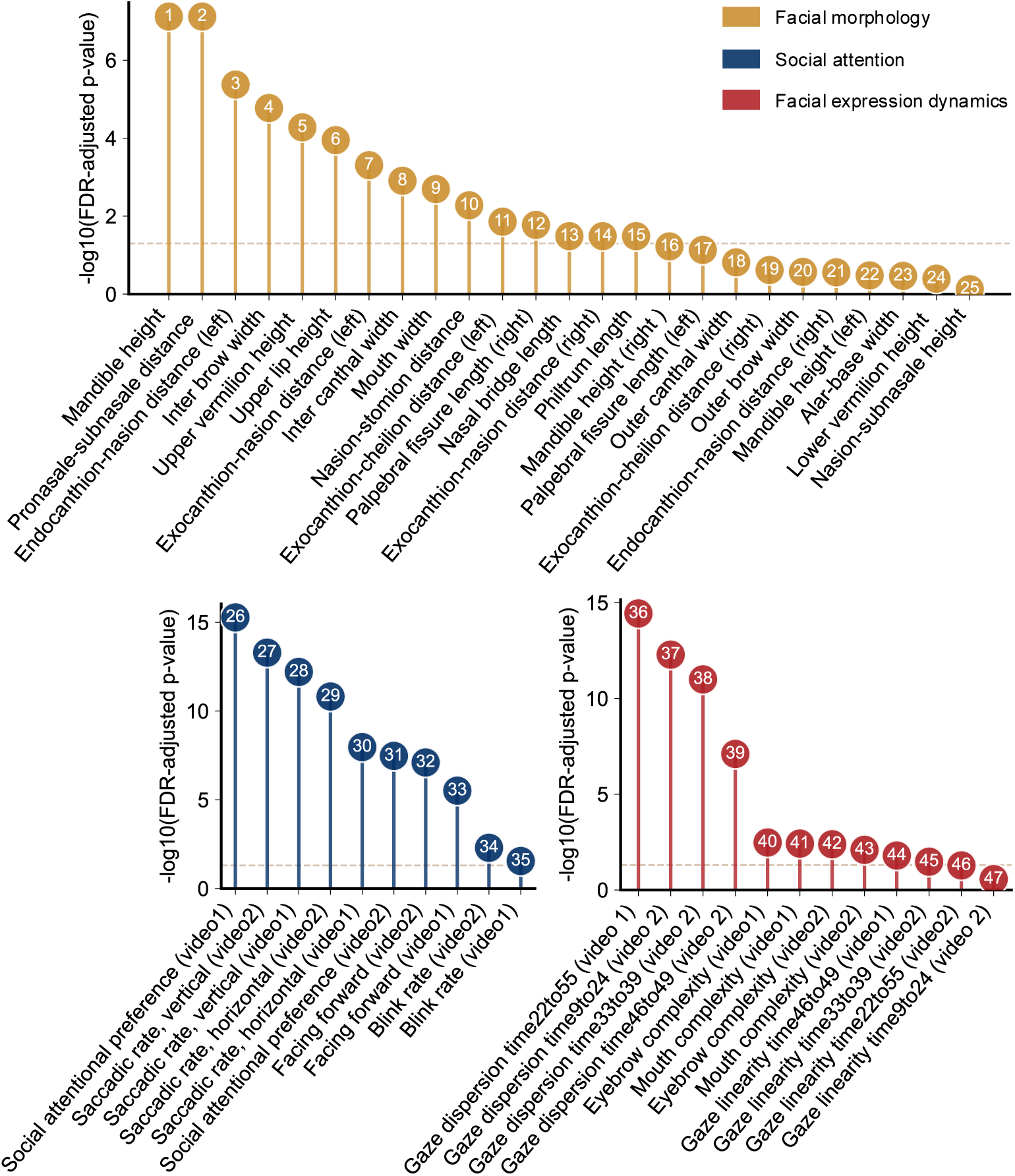
Identification of core digital behavioral and morphological phenotypes differentiating autism. A lollipop plot illustrating the statistical significance of differences in digital phenotypes between individuals with autism spectrum disorder (ASD) and typically developing (TD) controls. Phenotypes are categorized into three domains: facial expression dynamics, social attention patterns, and facial morphology. The highly significant differences found in key features, such as social attentional preference, gaze dispersion, and frequency of gaze changes, confirm that the collected data is rich with the quantifiable behavioral and morphological signatures of autism. Source data are provided as a Source Data file.

The most profound group differences were observed in social attention patterns. Gaze dynamics were a particularly strong differentiator; for example, social attentional preference (adjusted p *<* 10e-7 for both videos) and both vertical and horizontal saccadic rates were highly significant (all adjusted p *<* 10e-7). Gaze dispersion was also among the most significant discriminators across multiple video segments (e.g., adjusted p = 3.63e-15 and p = 5.15e-13). Furthermore, head orientation as a proxy for social engagement (i.e., “facing forward”) was a powerful differentiating phenotype (adjusted p *<* 10e-5 for both videos). Significant differences were also identified in static facial morphology. Key distinguishing features included mandible height (adjusted p = 7.63e-8), pronasale-subnasale distance (adjusted p = 7.63e-8), and inter-brow width (adjusted p = 1.69e-5), all of which surpassed the FDR significance threshold. In addition, we observed significant group differences in facial expression dynamics; for example, measures of complexity in eyebrow movements (eyebrow complexity; adjusted p *<* 0.005 for both videos) and mouth movements (mouth complexity; adjusted p = 3.79e-3) were highly significant.

Taken together, these results demonstrate that our video-based data collection protocol captures a rich set of quantifiable behaviors and static features that are distinct in ASD. This not only provides insight into the specific digital features that underpin the foundational AI model’s performance but also deepens our understanding of the characteristic behavioral signatures of autism.

### Identification of ASD subtypes and analysis using behavioral and clinical data

A critical goal of this study was to move beyond diagnosis and leverage the model’s learned representations to identify biologically informative ASD subtypes. Because the validation set was used for model selection, we merged it with the initial training set – treating the combined data as the final training set – to define the subtypes with maximal statistical power. Unsupervised clustering of the high-dimensional feature vectors extracted from the model’s internal layers revealed three distinct and robust ASD subtypes, hereafter referred to as Subtype 1, Subtype 2, and Subtype 3. The choice of three subtypes was determined empirically through a formal stability analysis, where a three-cluster solution (k=3) demonstrated the highest prediction strength (0.59) compared to other configurations (k=2, 4, 5), indicating it as the most stable and reproducible grouping within our data.

We then characterized these AI-defined subtypes by examining their unique digital behavior patterns across both training and discovery sets. Compared with TD, Subtype 1 (combined digital behavioral atypicality) exhibited the greatest derivation in both social attention patterns and facial morphology features; Subtype 3 (social attention atypicality) showed consistent divergence primarily in social attention patterns; while Subtype 2 (mild digital behavioral atypicality) displayed the least deviation, closely resembling the TD group. Fig. 4a illustrates how the three subtypes separate across representative digital phenotypes, with these differences replicating consistently across both training and discovery sets. The stability of these digital signatures in held-out test data underscores the robustness of the identified subtypes.

**Fig. 4:**
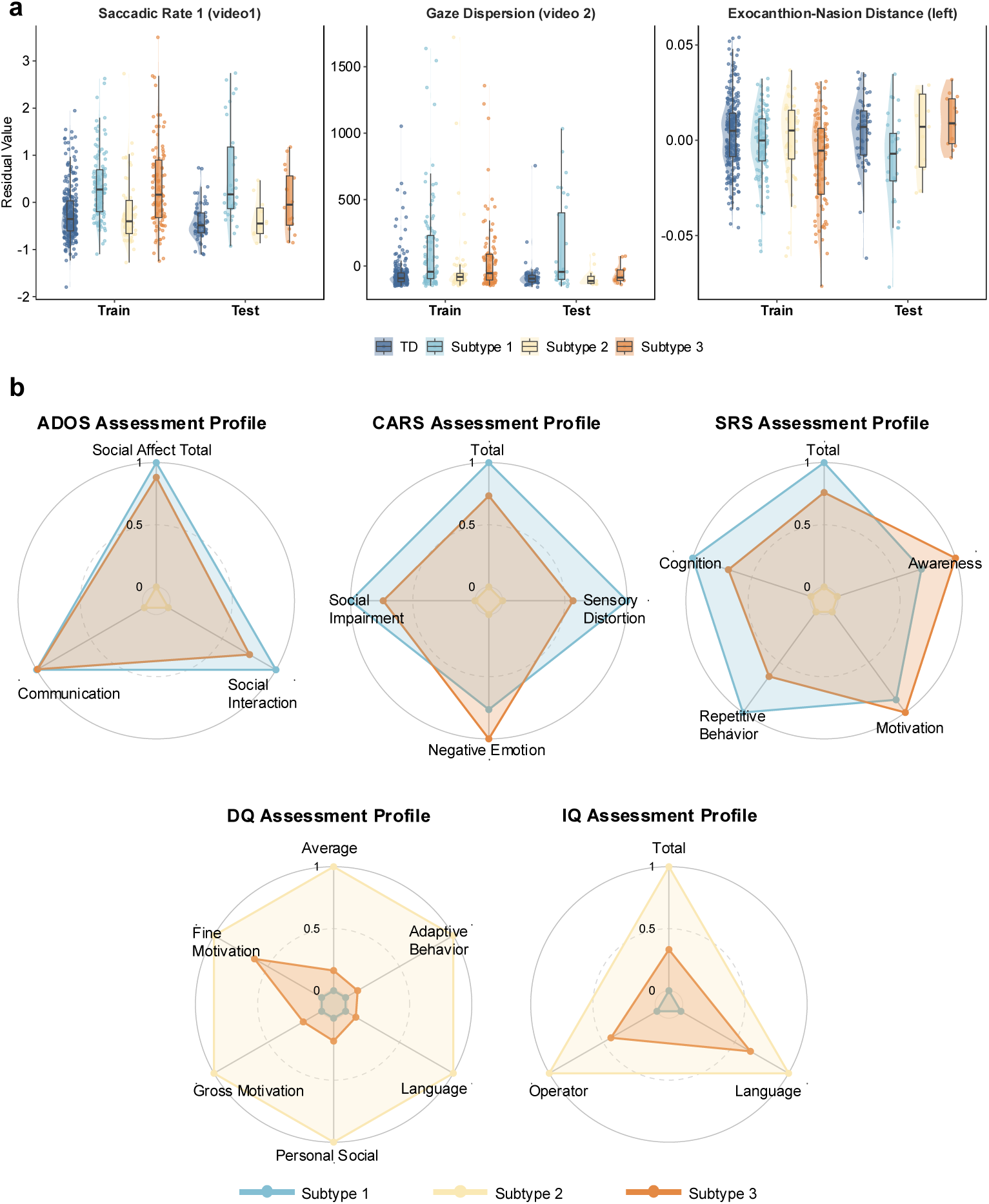
Foundational AI models identify three robust and clinically distinct subtypes of autism. **a**, Raincloud plots illustrating the distinct distributions of three representative digital phenotypes — saccadic rate, gaze dispersion, and exocanthionnasion distance — across the three identified ASD subtypes (Subtype 1, 2, 3) and the typically developing (TD) control group. The distinct patterns for each subtype are consistently replicated in both the train and discovery sets, demonstrating the robustness of the subtypes. **b**, Radar plots revealing the unique clinical profiles of the three ASD subtypes across four standard clinical assessments: the Autism Diagnostic Observation Schedule (ADOS), Childhood Autism Rating Scale (CARS), Social Responsiveness Scale (SRS), and developmental (DQ) and intelligence (IQ) quotient. Each axis represents a normalized subscale score, and the colored polygons represent the mean profile for each subtype. The distinct shapes of the polygons indicate that each subtype possesses a unique signature of clinical severity and behavioral characteristics. Source data are provided as a Source Data file.

To delineate the clinical and biological significance of these AI-derived subtypes, we mapped their profiles across a comprehensive battery of assessments, including the ADOS, CARS, SRS, and developmental (DQ) and intelligence (IQ) quotient assessments. As visualized in the radar plots (Fig. 4b), each subtype displayed a unique clinical signature. Subtype 1, which characterized by combined atypicalities in digital behavior, represents the most severe end of the spectrum. Individuals in this group demonstrated a global impairment profile, presenting with the most significant clinical symptoms across all domains of the ADOS, CARS, and SRS assessments. This profound clinical presentation was consistently mirrored by the lowest DQ and IQ across all measured domains. This suggests a widespread impact on foundational neurodevelopmental processes, affecting not only behavior but also core cognitive capacities. Conversely, Subtype 2, which exhibited only mild digital behavioral atypicality, aligns with a high-functioning clinical profile. This group presented with the mildest symptoms on all standardized clinical scales. Their adaptive capabilities were further evidenced by their superior cognitive and developmental profiles, consistently achieving the highest possible scores across all IQ and DQ domains. This suggests the presence of effective compensatory mechanisms or a less impactful neurobiological etiology, allowing for functioning that approaches neurotypical levels despite an ASD diagnosis. Subtype 3, presents the most nuanced profile, defined by specific atypicality in social attention. Clinically, they exhibited an intermediate level of impairment, with overall symptom severity falling between the other two groups. However, their profile was marked by a significant discrepancy: prominent challenges in specific areas, such as social communication on the ADOS, coexisted with less affected domains. This “spiky” profile of relative strengths and weaknesses was also reflected in their intermediate IQ and DQ scores. This specific pattern of social-attentional and communicative difficulty suggests a more targeted, rather than global, underlying neurobiological mechanism. The distinct challenges in this group highlight the critical need for interventions tailored specifically to social-cognitive processing, rather than a generalized approach.

### Analysis of brain structure patterns across ASD subtypes

To investigate the neurobiological underpinnings of the AI-derived subtypes, we examined whether they were associated with distinct patterns of brain morphometry using structural MRI data. After controlling for confounding variables such as age, sex, scanner, and the whole brain volume, we found significant neuroanatomical differences between the three subtypes, providing strong biological validation for their distinction.

As illustrated in Fig. 5a, these subtypes were associated with distinct patterns of cortical area, volume, and thickness, as well as subcortical volume. These differences were concentrated in brain regions critical for social cognition and sensory processing, including the lateral occipital, entorhinal, and inferior temporal cortices, and the temporal pole.

**Fig. 5:**
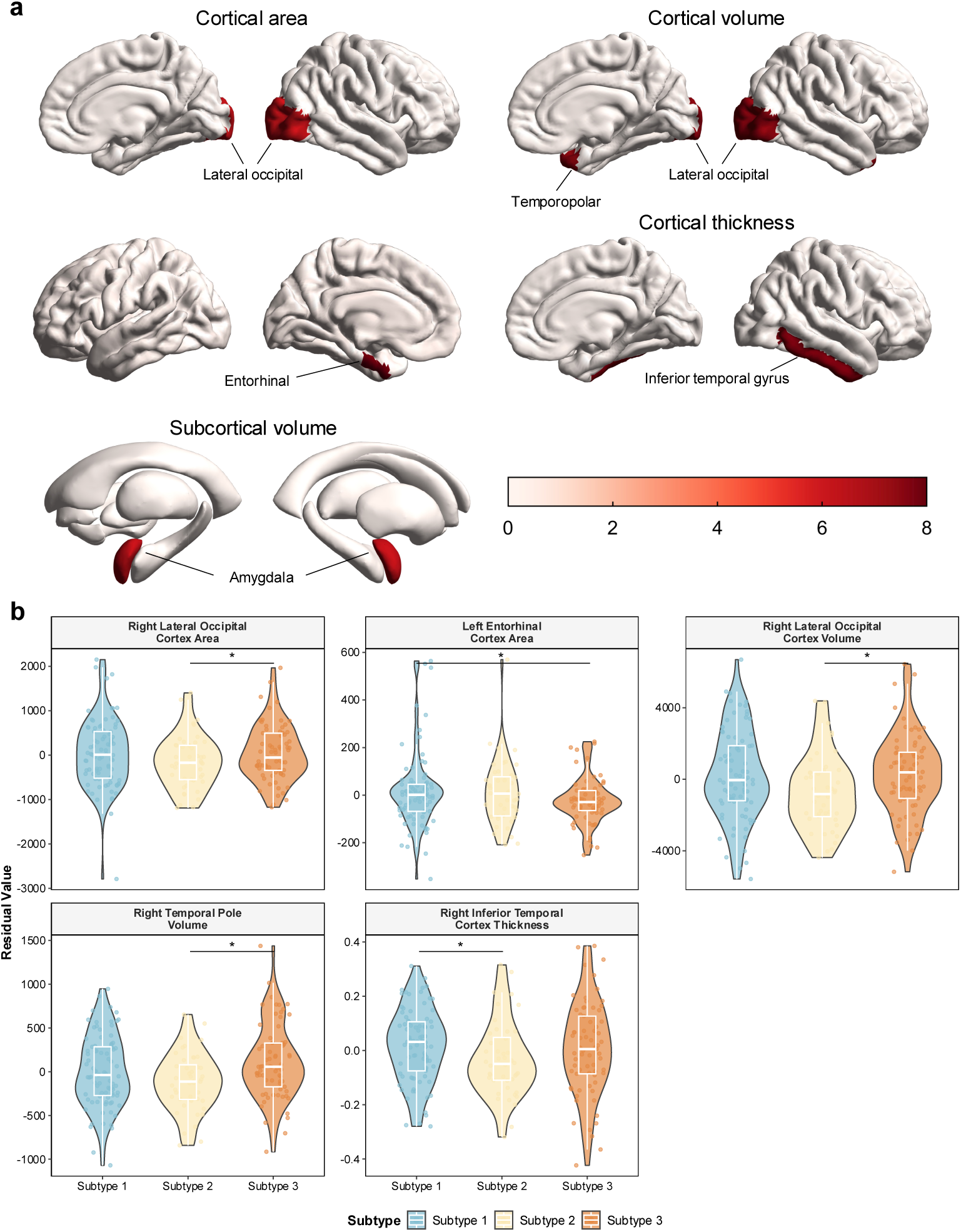
Neuroanatomical profiles of the three AI-derived ASD subtypes. **a**, Brain maps illustrating regions with significant structural differences among the three subtypes after controlling for age, sex, total brain volume and MRI scanners. Distinct patterns were observed in cortical area (lateral occipital, entorhinal), cortical volume (temporopolar, lateral occipital) and cortical thickness (inferior temporal gyrus). The color bar indicates the statistical significance of the differences. **b**, Violin plots showing the distribution of residual values for key structural metrics across the three subtypes. Significant between-group differences are highlighted (p *<* 0.05). Subtype 3 shows a significantly larger cortical area and volume in the right lateral occipital cortex compared to Subtype 2. Subtype 1 displays a larger cortical area in the left entorhinal cortex than Subtype 3 and greater cortical thickness in the right inferior temporal cortex than Subtype 2. These findings provide strong biological validation for the behaviorally defined subtypes. Source data are provided as a Source Data file.

Quantitative analysis confirmed that these anatomical differences aligned meaningfully with the subtypes’ behavioral and clinical profiles (Fig. 5b). Notably, several of the identified regions, such as the lateral occipital cortex and inferior temporal gyrus, are core components of the visual processing pathway. This finding is particularly relevant given the group differences observed in the gaze-based tasks. For instance, Subtype 3, which was characterized by highly atypical gaze patterns, showed a significantly larger cortical area and volume in the right lateral occipital cortex, and larger volume in right temporal pole compared to Subtype 2. This may underlie the distinct gaze patterns observed, since Subtype 2 had the least atypical gaze patterns while Subtype 3 showed worse gaze change and dispersion than Subtype 2 during the behavioral analysis. In addition, Subtype 1, which had the combined digital behavioral atypicality and most severe clinical profile, was distinguished by unique structural features, such as a larger cortical area in the entorhinal cortex compared to Subtype 3 and greater cortical thickness in the right inferior temporal cortex compared to Subtype 2, as well as trend difference in amygdala volume compared with Subtype 2 and 3.

Taken together, these findings provide compelling biological validation for our framework. They demonstrate that the AI-derived subtypes are anchored in distinct and functionally relevant neuroanatomical profiles. This convergence of evidence across behavior, AI-driven analysis, and neurobiology offers a robust, multi-modal approach to stratifying the autism spectrum.

## 3 Discussion

This study addressed two of the most pressing challenges in autism research: the need for scalable, objective diagnostic tools and the profound clinical heterogeneity that complicates treatment. Here, we demonstrate that a foundational AI model, requiring only two minutes of passive video viewing, can not only support rapid diagnostic screening but also uncover biologically meaningful subtypes within the autism spectrum.

Clinically, our work holds the potential to dramatically accelerate the diagnostic pathway for ASD. Current gold-standard assessments like the ADOS are resource-intensive bottlenecks, requiring lengthy administration by highly trained clinicians and limiting accessibility. While other AI tools have emerged, many still require 10 minutes or more of structured tasks, posing a barrier to seamless integration into busy clinical settings. Our two-minute protocol dramatically lowers this barrier, enabling feasible deployment in primary care or school settings, not just specialized centers. This accessibility is crucial for streamlining clinical workflows and reducing the often-protracted wait times for a formal diagnosis. The model’s high accuracy and robust performance across different ages, sexes, and cognitive levels confirm its reliability. Consequently, our method offers a powerful tool for rapid, large-scale risk stratification, facilitating earlier identification and faster access to vital early intervention services.

At the heart of our approach is a foundational AI model that learns end-to-end from raw video data, thereby minimizing human bias in feature engineering. Beyond a simple binary diagnosis, the model generates a continuous output probability score. It correlated with established clinical measures, including the ADOS, CARS, and SRS, validating that the AI captured patterns directly relevant to clinical assessments. Furthermore, to provide explainability, we correlated this same score with a rich set of independently quantified digital phenotypes. This confirmed that the model’s predictions were driven by clinically meaningful behaviors, including patterns in social attention and facial expression. Thus, the model provides not only a rapid prediction but also an objective, interpretable metric rooted in specific, quantifiable behaviors.

A central contribution of this work is the data-driven discovery of distinct ASD subtypes, directly addressing the challenge of clinical heterogeneity. By applying unsupervised clustering to the AI’s learned representations, we identified three stable and replicable subgroups. Crucially, these subtypes were not mere statistical artifacts; they were validated as clinically distinct, with clear differences in their behavioral profiles, from severe to mild presentations. This provides a novel, data-driven framework to stratify the autism spectrum, paving the way for future research into subtype-specific neurobiology and the development of targeted, personalized interventions.

We provided a critical anchor for these behavioral subtypes with neurobiological evidence. The AI-defined subgroups corresponded to distinct differences in brain structure. We found significant variations in cortical area, volume, and thickness in brain regions vital for social cognition and visual processing, such as the inferior temporal and lateral occipital cortices. This convergence of behavioral, clinical, and neurobiological data supports the biological validity of our subtypes and creates a powerful link between observable behavior and its underlying neural substrates.

Our findings should be considered in light of several limitations, which in turn define critical directions for future work. First, while the model performed well, it requires validation in larger, more diverse cohorts to ensure generalizability and equity. Second, the cross-sectional design of this study should be followed by longitudinal research to determine if these subtypes are stable over developmental periods and if they can predict differential responses to intervention. Finally, continued work with explainability techniques is needed to further illuminate the specific behavioral cues the foundational AI model uses for its predictions.

This work establishes a scalable, objective framework for both clinical stratification and diagnostic support in autism. By linking brief behavioral recordings to distinct clinical and neurobiological profiles, we offer a path towards deconstructing the heterogeneity of ASD. Such an approach opens new avenues for research into targeted therapies and personalized care. It represents a foundational step from a “one-size-fits-all” view of the spectrum towards a deeper understanding that can guide more effective, individualized support for every child with autism.

## 4 Methods

### 4.1 Study participants

The used dataset comprised videos of children watching standardized video content, including both children diagnosed with ASD and Typically Developing (TD) children. The diagnosis of ASD was established according to the Diagnostic and Statistical Manual of Mental Disorders, Fifth Edition, then confirmed using the Autism Diagnostic Observation Schedule, Second Edition (ADOS-2). Children with ASD were also assessed with Childhood Autism Rating Scale (CARS) and Social Responsiveness Scale, Second Edition (SRS-2). Demographic data, including age and gender, were collected for all participants. Children with ASD were enrolled from the Department of Developmental Behavior Pediatrics, Shanghai Jiao University School of Medicine Affiliated Xinhua Hospital; TD children were recruited during routine community health care. Inclusion criteria: Children aged 15 months and older were included. ASD diagnoses for children enrolled under 24 months of age were reconfirmed at 24 months. Exclusion criteria: Individuals diagnosed with a known genetic disorder, and those with craniofacial malformations potentially caused by a genetic disorder (evaluated by clinical geneticist); Severe sensory (visual or auditory) or motor impairments; Uncontrolled epilepsy; Acute illness precluding valid data collection; Inability or unwillingness to sit on a parent’s lap while viewing the stimulus video. The dataset was randomly partitioned into training, validation, and testing sets, with 60% allocated to training, 20% to validation, and 20% to testing. An external validation set was obtained from the Department of Child Healthcare, Huaibei Maternal and Child Health Hospital, Early Life Plan Alliance of Xinhua Hospital. This study protocol was approved by the Ethical Committee of Xinhua Hospital (XHEC-C-2024-085-1) and the Ethical Committee of Huaibei Maternal and Child Health Hospital (FYL2024014), and legal guardians gave written informed consent for participants.

### 4.2 Video content and data collection

We devise two videos that contain different social stimulus. The first video (67-second) assesses social preference. An adult male is on one side of the screen, and the bubbles he blows are on the other. A distractor is a toy car in the top corner on the bubbles’ side. The second video (51-second) assesses joint attention, divided into the responding joint attention (RJA) section and the initiating joint attention (IJA) sections. In the RJA section, an adult female turns to a toy brick and stares at it, prompting the child to look at the same brick. In the IJA section, the female looks straight ahead. Two toy cars appear on either side; one moves forward, the other disappears then reappears. This checks if the child will share interest with the female through eye contact. For more detailed information about these videos, please refer to Supplementary Videos 1 and 2.

The child either sit independently or is seated on the caregivers’ laps as instructed. A mobile phone, fixed on a holder, is placed 30 centimeters away from the child. Then, two short videos are played on the phone screen. As the child watches the videos, the frontal camera embedded in the device records the child’s behaviors, at resolutions of 640*×*480, 25 frames per second. The caregivers are requested to refrain from instructing their child. Data collection was conducted in a quiet room with uniform lighting to minimize environmental confounds.

### 4.3 ASD prediction with AI foundational model

ASD prediction from behavioral video data is a complex task, characterized by subtle cues and significant inter-individual variability. The conventional reliance on labor-intensive feature engineering might be misleading, as it injects implicit biases and prior assumptions into the model, and discards potentially valuable information present in the raw video data. To overcome these limitations, we formulate ASD prediction as an end-to-end video classification problem and leverage VideoMAE [16], a video foundational model pre-trained on large-scale video datasets. VideoMAE, inspired by masked autoencoding techniques in natural language processing and image analysis, is a self-supervised video pre-training framework that learns robust video representations by reconstructing masked spatiotemporal “tubes” within video clips. We use VideoMAE-base pre-trained on Kinetics-400 [24] (240,000 training videos; 400 human action classes) as the backbone for feature extraction. This large-scale pre-training equips the model with generalizable spatiotemporal representations, which we adapt to ASD prediction task through fine-tuning on our dataset of children’s watching videos. These videos capture children’s naturalistic behavioral responses, from which the model learns to distinguish between ASD and non-ASD cases.

The fine-tuning process involves optimizing the model’s weights using standard supervised learning techniques. We adopt the official VideoMAE implementation and pre-trained weights available through HuggingFace. Default hyperparameters are retained unless the ones in bold that we changed to account for the large sample size and the parameter of the recorded videos, and to accelerate training. **batch size** = **64** instead of 8 to accelerate training; num frames to sample = 16; sample rate = 4; **fps** = **25** instead of 30 due to the parameter of the recorded videos; **num epochs** = **8** instead of 4 due to the large sample size; learning rate = 5*e −* 5; warmup ratio = 0.1.

### 4.4 Statistical analysis

Model training was performed using Python (V.3.12.4). The deep learning infrastructure utilized PyTorch (V.2.3.1) as the core framework, augmented by PyTorchVideo (V.0.1.5) for video data handling and the transformers library (V.4.44.0) for Video-MAE implementation.

For performance evaluation, the dataset was divided into three distinct subsets: a training set for model optimization, a validation set for model selection, and an independent test set for final assessment. This tripartite split ensures that the model’s predictive capabilities are assessed on data not used during training or tuning, providing an unbiased estimate of its generalizability. The Receiver Operating Characteristic Area Under the Curve (ROC AUC) was adopted as the primary performance metric to measure the model’s ability to distinguish between classes across all classification thresholds. For all metrics — including AUC, sensitivity, specificity, Positive Predictive Value (PPV), and Negative Predictive Value (NPV) — 95% Confidence Intervals (CIs) were derived using the percentile bootstrap method with 2,000 resamples. For threshold-dependent metrics (sensitivity, specificity, etc.), the operating point was determined for each bootstrap sample by finding the threshold that maximized the Youden index (sensitivity + specificity - 1). This approach accounts for uncertainty in both the sample data and the selection of the optimal threshold.

Beyond classification, we sought to quantitatively validate the model’s continuous output as a clinically meaningful measure. We then evaluated the relationship between the model-derived probability and a comprehensive set of clinical assessment scores (including CARS, ADOS, and SRS total and subscale scores) on a cohort of 243 participants diagnosed with ASD. The association between the model-derived probability and clinical measures was assessed using two complementary statistical methods. First, a series of simple linear regression analyses were performed to determine the extent to which the model-derived probability (independent variable) could predict scores on each clinical measure (dependent variables). For each regression, we calculated the standardized beta (*β*) coefficient, the coefficient of determination (R^2^), and the corresponding p-value. Second, to ensure that observed associations were not driven by potential confounding factors, a partial correlation analysis was conducted. This method assessed the direct relationship between the model-derived probability and each clinical measure while statistically controlling for the participant’s age, sex, and developmental/intelligence quotient (IQ/DQ), providing the partial correlation coefficient (r) and its p-value.

Statistical analyses, including the computation of performance metrics and the correlational and stratified analyses, were conducted using the SciPy (V.1.14.1) and scikit-learn (V.1.5.1) libraries. A significance level of p*<*0.05 was used for all statistical tests.

### 4.5 Computation of subtypes

To identify data-driven behavioral subtypes, we developed and applied an unsupervised machine learning pipeline to the cohort’s behavioral videos. This pipeline systematically transformed high-resolution video data into low-dimensional representations suitable for clustering. The process involved three core stages: deep feature extraction, subject-level feature aggregation and dimensionality reduction, and finally, clustering to define subtypes.

We first extracted rich, high-level behavioral features from each participant’s two video recordings. This was accomplished using a foundational AI model (i.e., Video-MAE), which had been specifically fine-tuned for ASD classification to sensitize it to clinically relevant behavioral patterns. Each video was processed as a sequence of short, non-overlapping clips. The fine-tuned model transformed each clip into a high-dimensional feature vector, effectively converting each participant’s raw video data into a temporal sequence of learned representations.

To generate a single, comprehensive feature vector for each participant, all clip-level representations derived from that child’s videos were concatenated into one long-form vector. The resulting high-dimensional subject representations were then reduced using Non-negative Matrix Factorization (NMF) [25, 26]. NMF is an ideal technique for this task as it learns a parts-based, additive representation of the data, which can enhance the interpretability of the resulting subtypes. We applied NMF to the cohort’s feature matrix to learn 32 latent components. This process yielded a compact, 32-dimensional feature vector for each participant, capturing the most salient behavioral information. The 32-dimensional NMF feature vectors were used to partition the ASD cohort into subtypes using the k-means algorithm. To maximize the data available for discovering robust clusters, we first merged the training and validation sets into a single training cohort. The optimal number of clusters (k) was then determined on this merged cohort using the prediction strength method [27], a formal statistical approach that measures cluster stability across data splits. Prediction strength is particularly well-suited for assessing the generalizability of the clusters to new data, a critical aspect for clinical applications [28]. We evaluated k from 2 to 5, and this analysis revealed a clear peak in prediction strength at k = 3 (strength = 0.59), indicating it as the most stable and reproducible clustering solution for our data. With k = 3 established, clustering was performed separately on the merged training cohort and the held-out discovery set. To resolve the arbitrary label assignments from these independent runs, we employed bipartite graph matching using the Hungarian algorithm [29]. This created a one-to-one mapping between the clusters from both sets by maximizing their subject overlap. The final output was three matched and reproducible behavioral subtypes, which were carried forward for multi-modal validation.

### 4.6 Computation of phenotyping

To facilitate the interpretation of the trained classification model and understand the relationship between its learned representations and clinically relevant or observable characteristics, we extracted features from multiple modalities. These features were categorized into five distinct groups derived from video recordings and structural neuroimaging, complementing the model-derived features (predicted probability, implicit embedding), clinical assessments (ADOS, ABC, SRS), and demographic data (age, gender) used in subsequent analyses.

#### Saccade

This group of features quantifies aspects of eye movement dynamics and gaze stability and contains three types of features. The first one is gaze change intensity. We estimated the intensity of gaze shifts using frame-by-frame 2D gaze direction vectors obtained from the L2CS-Net model [30]. The Euclidean distance (L2 norm) between consecutive gaze vectors was calculated for each frame transition, providing a measure of gaze velocity or change intensity. The average intensity was used as the feature value for each participant. The second one is facing forward. To approximate the child’s attention towards the screen stimuli, we calculated the proportion of time they were “facing forward”. Using facial landmark and head pose data extracted by OpenFace (V.2.2.0), frames were filtered based on three criteria: (i) eyes detected as open, (ii) estimated gaze vector directed at or near a predefined screen area, and (iii) relatively stable head pose [7, 31]. The percentage of frames meeting all criteria constituted the facing forward feature. The third one is blink rate. Eye blinks, often associated with attentional engagement and cognitive load, were detected using OpenFace based on action unit detection (AU45 is used here). The total number of blinks detected during valid viewing periods (*e.g.*, when facing forward) was normalized by the duration of these periods to compute the blink rate.

#### Social attention

This group assesses how children allocated their visual attention to specific spatial locations or dynamic events within the video stimuli. This group contains three types of features. The first is gaze landing point distribution. Gaze landing points relative to the screen were derived using the iTracker model [32]. We quantified the spatial distribution of gaze fixations for specific videos. For Video 1, the proportion of valid gaze points landing on the left versus the right halves of the screen was calculated. For Video 2, the proportion of gaze points landing on the top versus the bottom halves was determined. These proportions reflect potential biases in spatial attention allocation. The second is gaze linearity. Specific segments within the stimuli involved predictable motion paths (*e.g.*, a car moving linearly in Video 1). To assess how closely a child’s gaze followed these paths, we applied Principal Component Analysis (PCA) to the sequence of 2D gaze coordinates recorded during these segments. The explained variance ratio of the first principal component served as a measure of gaze linearity, with higher values indicating gaze patterns more concentrated along a single axis. The third is gaze dispersion. As a complementary measure to linearity, gaze dispersion quantified the overall spread of gaze points during the same stimulus segments. This was calculated from the covariance matrix of the 2D gaze coordinates. Specifically, we used the trace (sum of eigenvalues) of the covariance matrix as an indicator of the total variance of the gaze points during the segment. Higher values indicate greater dispersion.

#### Facial dynamics complexity

To capture the richness and variability of facial expressions, we measured the complexity of facial movements in key social signalling regions. This group contains two types of features: eyebrow and mouth complexity. Time series data representing the movement of facial landmarks (extracted via Open-Face) corresponding to the eyebrow and mouth regions were generated. We applied multiscale entropy analysis [33] to these time series data to quantify their complexity across multiple time scales. Integrated MSE values were computed separately for the eyebrow and mouth regions.

#### Facial morphology

Static facial morphology features were quantified to explore potential associations between facial structure and ASD characteristics or model predictions. We utilized the 2D facial landmarks extracted by OpenFace. From these landmarks, we calculated a series of specific distances in pixels between designated landmark pairs. To account for variations in apparent face size due to head size differences and distance from the camera, each measured distance was normalized by a reference facial width. This reference width was defined as the Euclidean distance in pixels between landmark 0 and landmark 16, which correspond approximately to the left and right edges of the jawline/face outline in the OpenFace landmark scheme. This measurement was selected because these landmarks represent the widest part of the face (bizygomatic width) and provide a stable, anatomically consistent measure of facial width, as they are located at the fixed bony structures of the zygomatic arches and are less affected by facial expressions or soft tissue movements compared to other facial landmarks [34, 35]. We computed 25 distinct facial morphology features, each representing the ratio of a specific anatomical distance to this reference width. These features included ratios corresponding to standard anthropometric measurements such as outer canthal width (distance between landmarks 45 and 36, normalized), alar base width (landmarks 34 vs. 32, normalized), nasal bridge length (landmarks 27 vs. 30, normalized), upper lip height (landmarks 33 vs. 62, normalized), inter-brow width (landmarks 21 vs. 22, normalized), mouth width (landmarks 48 vs. 54, normalized), and others capturing dimensions of the eyes, nose, mouth, cheeks, and overall facial proportions. The complete list of the 25 calculated ratios and their corresponding landmark pairs is detailed in Supplementary Table 1. For each participant, the final feature value for each ratio was typically computed as the average value obtained from the selected facing forward frames.

#### Neuroimaging

Structural MRI data were acquired using either a Siemens Verio 3.0T or Philips Ingenia 3.0T scanner, both equipped with a 32-channel head coil and 4-channel neck coil. High-resolution T1-weighted magnetization-prepared rapid gradient-echo (MPRAGE) images were obtained for anatomical reconstruction. T1-weighted images were processed using FreeSurfer V.7.3.2. Morphometric measures consisting of the cortical area, volume, thickness, curvature, and folding index of 68 cortical regions(parcellated via the Desikan-Killiany atlas) and volume of 40 sub-cortical regions (via aseg atlas). All T1-weighted images were visually inspected for quality and motion artifacts prior to processing. Participants with excessive motion or anatomical abnormalities were excluded from the neuroimaging analysis.

## Declarations

### Ethics approval and consent to participate

The Ethics Committee of Xinhua Hospital, Shanghai Jiao Tong University School of Medicine, approved this work (XHEC-C-2024-085-1). In adherence to the Declaration of Helsinki, written informed consent was provided by the parents or legal guardians of each participant.

### Consent for publication

Not applicable.

### Data availability

The data supporting this study are subject to strict confidentiality requirements. Access is restricted to authorized institutions under a formal data sharing agreement and ethical approval.

### Materials availability

Not applicable.

### Code availability

The source code and implementation details of this study are protected by intellectual property rights and are not publicly available. Access is restricted to authorized institutions under a formal data use and licensing agreement. Key components of the code have been provided in the supplementary materials for review purposes.

### Author contribution

F.L. and W.W. supervised the research. F.L. and W.W. initiated and conceived the main concept of the work. W.W. provided the key AI idea. W.W. and G.C. developed the AI methodology, wrote the code, and performed the experiments. W.W., F.L., G.C., W.Z., L.Z., Y.J., and Y.W. interpreted the results. X.H., L.Z., Y.J., Q.Z., T.R., H.T., J.C., K.L., X.S., S.H., L.G., J.L., H.W., and G.S. participated in the video data collection. All authors wrote and approved the final manuscript.

